# The Efficacy of Multimodal Physiotherapy versus Usual Care in Chronic Whiplash-Associated Disorders With Facet-Mediated Pain Undergoing Platelet Rich Plasma treatment: A Series of Single Case Experimental Designs

**DOI:** 10.1101/2023.10.16.23296769

**Authors:** Ashley Smith, Arun Gupta, Jacqui Stone, Jeff Habberfield, Geoff Schneider

## Abstract

**Purpose:** Chronic whiplash-associated disorders (WAD) is a heterogeneous condition with limited effective treatment options available. This study evaluated the effectiveness of multimodal physiotherapy versus usual care in chronic WAD with facet-mediated pain receiving platelet-rich plasma (PRP) injections to determine if pain interference and confidence completing activities in the presence of neck pain could be improved.

**Materials and Methods:** A multiple-baseline, single-case experimental design was used to evaluate the 6-week effect of physiotherapy or usual care in two groups of three participants each.

**Results:** All six participants demonstrated a significant reduction in pain interference, and three participants showed improved confidence to perform daily activities when in pain. Weighted Tau-U demonstrated a significant reduction of pain interference with large to very large effect sizes (> 0.75) for both interventions in all participants irrespective of treatment allocation with no significant group difference demonstrated. Similar effects were demonstrated for the confidence to perform daily activities with neck pain (ES > 0.46), although this was only evident in three participants (two PT and one UC). Generalization measures also showed improvements in pain and disability, psychological and quality of life outcomes. No adverse events were reported.

**Conclusions:** Both physiotherapy and usual care demonstrated improvements in pain interference and confidence to perform activities of daily living with neck pain in people with chronic WAD following cervical facet joint PRP.

**Trial Registration:** The trial was registered with ClinicalTrials.gov (Protocol Number: NCT03949959)

## Introduction

Up to 50% of people who experience a whiplash injury continue to report symptoms 12-months later (Carroll et al., 2008), with approximately 20% still recalcitrant to treatment 2-years post-injury (Kamper et al., 2008). This is problematic as very few treatments for chronic whiplash-associated disorders (WAD) demonstrate effectiveness in reducing pain and disability levels (Shearer et al., 2015; Teasell et al., 2010).

Physiotherapy is recommended in management guidelines for chronic WAD (Blanpied et al., 2017; TRACsa, 2008), but evidence is lacking regarding its effectiveness (Sutton et al., 2014). Recent trials only demonstrate a modest benefit of physiotherapy when analyzed at the group level (Jull et al., 2007; Ludvigsson et al., 2015; Michaleff et al., 2014; Ris et al., 2016; Stewart et al., 2007). Furthermore, an observational study demonstrated that approximately 50% of people with chronic WAD attended multidisciplinary treatment, inclusive of extensive multimodal physiotherapy, without improvement in health outcomes (Smith et al., 2013). However, following a pain-relieving intervention (cervical radiofrequency coagulation – RFC) directed at the underlying tissue lesion (cervical facet joints), health outcomes significantly improved (Smith et al., 2015). It must be noted in this study that participants were able to pursue concurrent conservative rehabilitation. Another recent observational case series also demonstrated significant improvement in pain and disability levels following cervical facet joint platelet-rich plasma (PRP) injections in people with chronic WAD (Smith et al., 2022). Many participants in this study also attended conservative rehabilitation following the intervention. Other studies investigating the effects of interventional therapies do not document whether participant’s attend subsequent physical rehabilitation (Barnsley, 2005; Lord et al., 1996). Hence, it is unclear if the reduction in pain and disability observed following these medical interventions are primarily resulting from the interventions themselves, subsequent rehabilitation, or a combination of the two.

Thus, in this proof-of-concept study, we investigated a standardized evidence-based multimodal physiotherapy intervention (Jull et al., 2008) in people with chronic WAD and confirmed facet-mediated pain undergoing cervical facet joint PRP and compared health outcomes to people undergoing cervical facet joint PRP with usual care (UC). Single Case Experimental Designs (SCEDs) were used to investigate the research hypotheses. In contrast to an experimental group design in which one group is compared with another at the group level, participants in single-subject research provide their own control data for the purpose of comparison in a within-subject rather than a between-subjects design. Additionally, single cases can be aggregated to arrive at a group effect.

## Methods

The study investigated a series of multiple baseline single case experimental designs (SCEDs) in chronic whiplash-associated disorders (WAD) comparing the effect of multimodal physiotherapy (PT) in people undergoing Platelet-Rich Plasma (PRP) versus PRP with usual care (UC). The efficacy of the intervention was evaluated using a multiple-baseline design (MBD) across participants (Lillie et al., 2011; Nikles, 2015).

### Design

In the context of sample size and sampling frequency, the study design was based on single-subject research design guidelines (Romeiser Logan et al., 2008). To evaluate the efficacy of the intervention (PT) it is recommended that a minimum of three participants and five data collection points in each phase are included in the study design (Romeiser Logan et al., 2008). This study utilized an A-B design across two phases: a baseline (A: no active or control treatment provided) and intervention phase (B: PT or UC). Participants in the MBD – to enhance internal validity – were either randomized into PT or UC and blinded to treatment allocation. Participants completed outcome measures for target behaviours into a daily paper diary across both the baseline and intervention phases. Participants in each treatment allocation were also randomized (and blinded) into a variable length baseline (A) with a staggered start – one participant in PT/UC starting at 5 days (post-PRP), one at 8 (post-PRP) and one at 11 days post-PRP (Fig. 1). Randomization was performed by one of the study authors (GS) using a computerized random number generator (random.org). The intervention phase was followed by a 12-week follow-up phase where participants did not have planned contact with the intervention personnel. This follow-up phase was implemented to determine the possible duration of improvement post intervention. Secondary outcome generalization measures for this period were only recorded at the end of the follow-up period. The daily diary patient reported outcomes were collected every two weeks during the intervention phase by the treating physiotherapist or study co-ordinator and passed to the blinded outcome assessor for data entry. The study was conducted and reported according to the Single-Case Reporting guideline in Behavioral interventions (SCRIBE) 2016 Statement (Tate & Perdices) (Supplementary Information) and registered through ClinicalTrials.gov (Protocol Number: NCT03949959). Institutional ethical approval was received for this study (REB18-0724), based on the submitted trial protocol (available from the corresponding author) and informed consent was obtained from all participants before enrolment.

**Figure 1:**
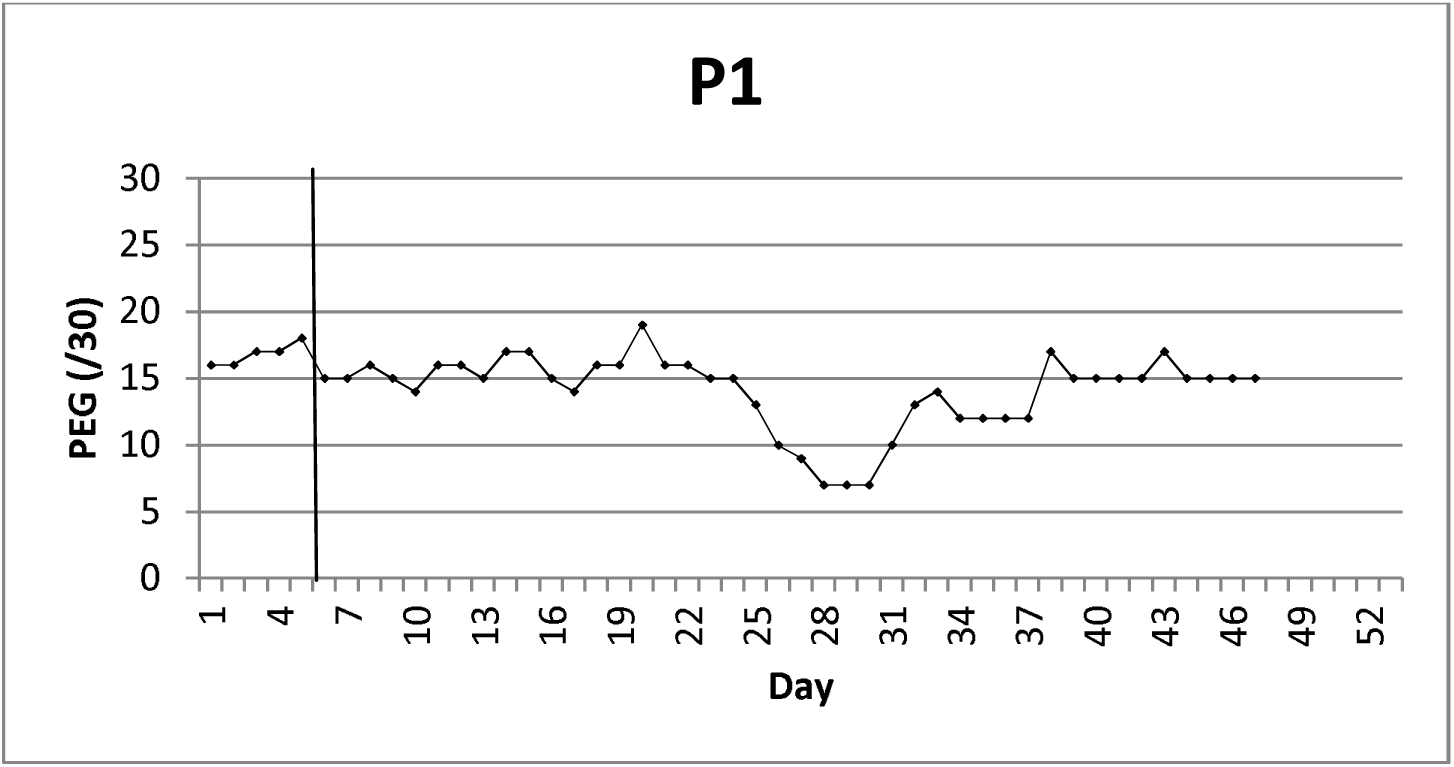
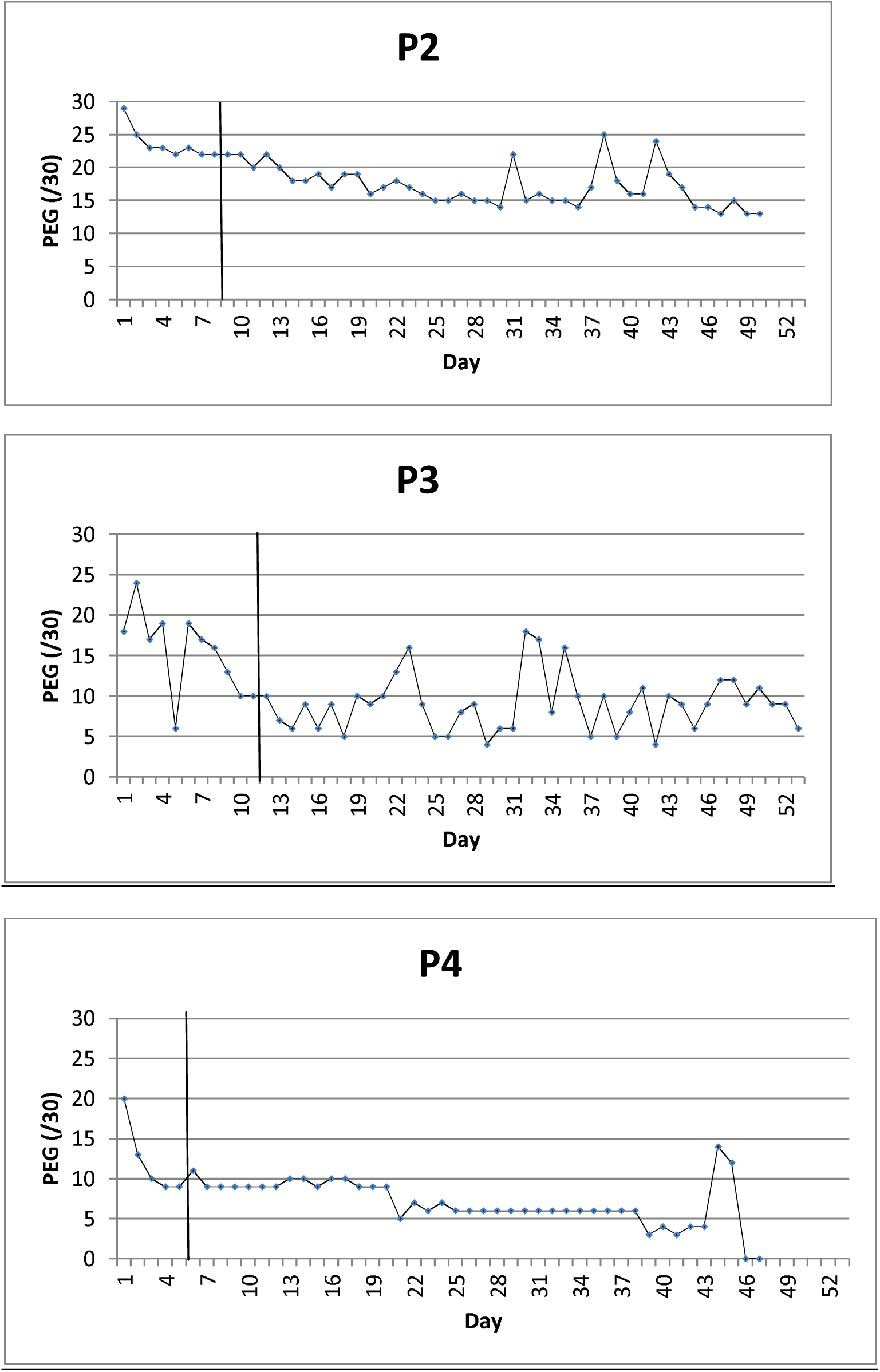
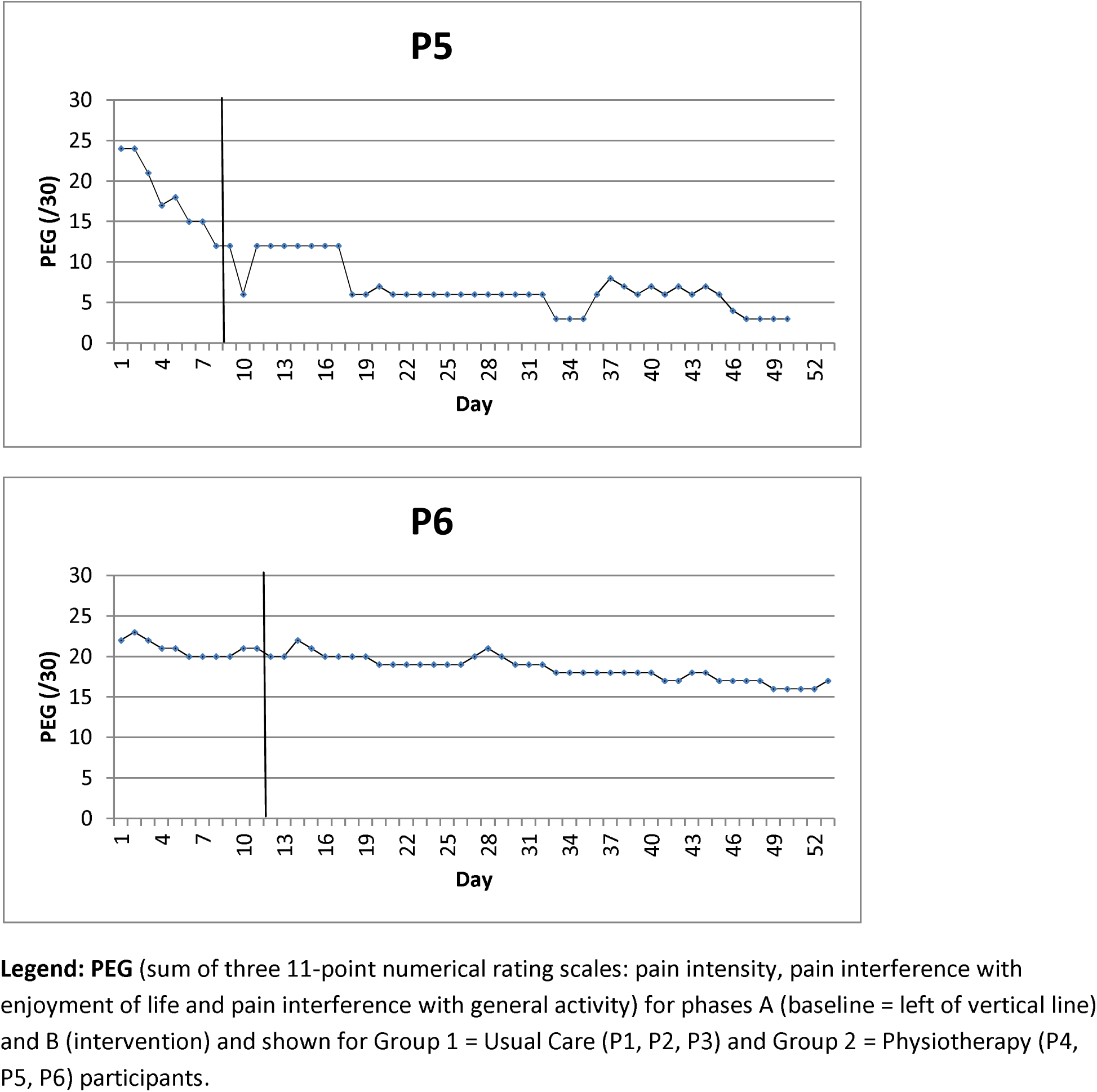
A two section AB graph plotting PEG scores of six people undergoing PRP - three receiving usual care and three receiving physiotherapy.

### Participants

Three participants were enrolled in UC (Participants 1, 2, 3) and three enrolled in PT (Participants 4, 5, 6). As all participants received PRP, this also allowed for replication of the study to examine external validity and for generalization of the effect of PRP (Kazdin, 2011). Participants were eligible to participate if they fulfilled all inclusion and exclusion criteria (Table 1).

**Table 1:**
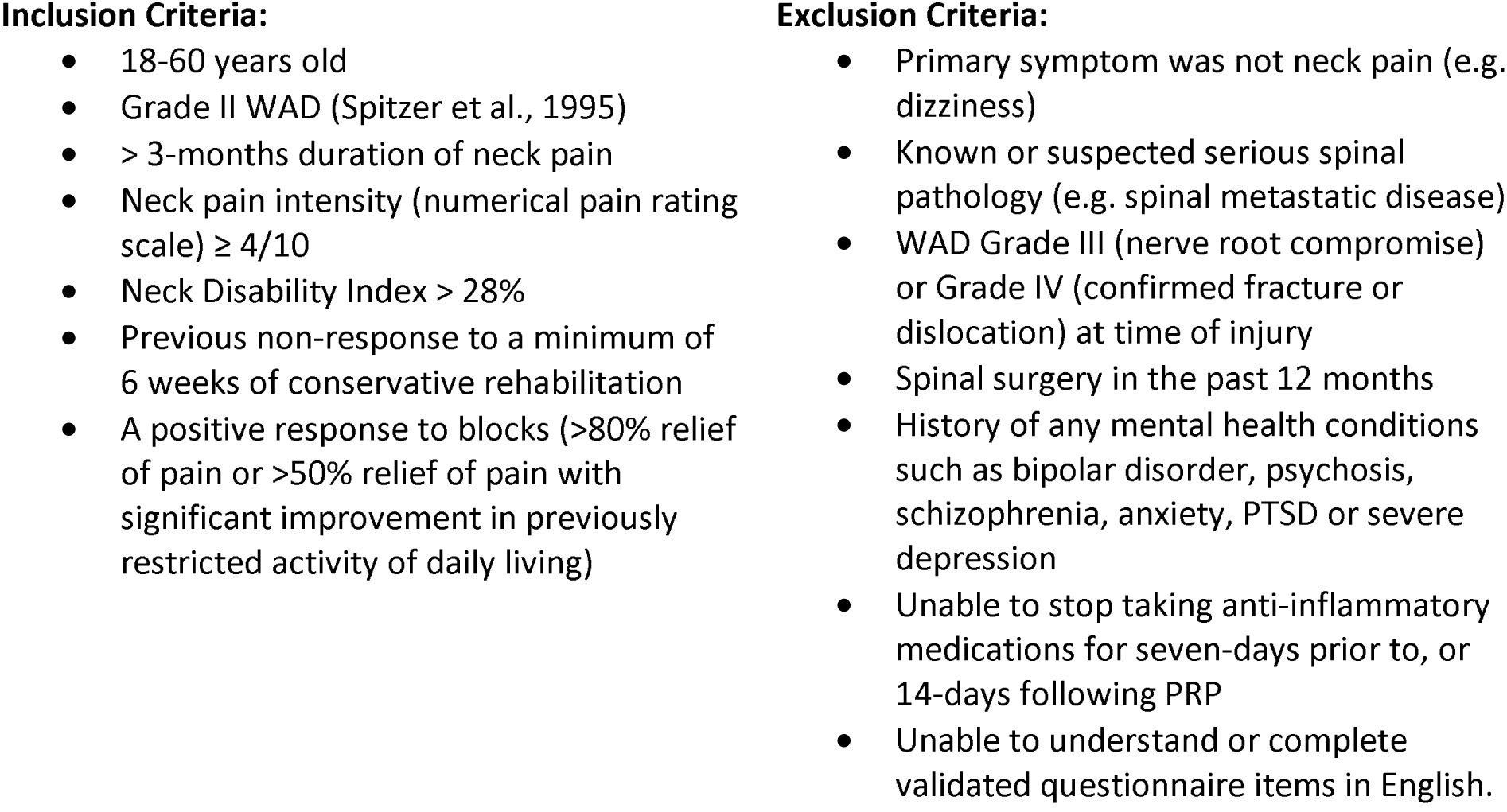
Inclusion and Exclusion Criteria.

### Interventions

All participants received PRP injections into the putative cervical facets and an educational pamphlet and were randomized into UC or PT (Appendix: Table 1).

#### Group 1 - Usual Care/Informational Booklet

Participants randomized to six-weeks of usual care (UC) were provided with an advice booklet Whiplash Injury Recovery: A Self Help Guide (3^rd^ edition) (Queensland et al., 2019) and education regarding PRP and associated healing cycles (Appendix: Table 1).

#### Group 2 - Multimodal Physiotherapy

As per established clinical practice guidelines linked to the International Classification of Functioning, Disability and Health (From the Orthopaedic Section of the American Physical Therapy Association) (Blanpied et al., 2017), multimodal physiotherapy was provided for six weeks, which consisted of advice, education, reassurance, manual therapy, intramuscular needle interventions and goal-directed exercise progressions (Appendix: Table 1).

At the end of the trial, in respect of the compensable nature of a whiplash injury, the participants were permitted to seek further treatment if required. Information about any additional treatments sought by participants (e.g. additional medication, physiotherapy etc.) was gained via patient diaries at the follow-up time points.

### Study Personnel

An experienced musculoskeletal physiotherapist with doctoral level postgraduate qualifications provided the therapeutic interventions for this study. The physiotherapy exercise sessions were audited twice (at completion of week two and week four) to check for adherence to the exercise protocol. A separate assessor blinded to study treatment allocation was responsible for collecting and entering the primary and secondary outcome data.

### Outcome Measures/Target Behaviours

Outcome measures for target behaviours were associated with pain reduction as it related to function – less pain interference in general activities and enjoyment of life and confidence to participate in daily activities despite pain. Repeated measures of the target behaviours were used to address individual variability. It is recommended that five data collection points are collected in each phase to evaluate a SCED intervention (Kratochwill et al., 2013). Target behaviours were evaluated prior to administration of PRP and then daily in the baseline phase during the first 5-11 days following PRP, with further daily measurements collected over the ensuing 42 days in the post-intervention phase. Participants completed these measures in a daily pain diary. These were forwarded to the research staff at the end of the baseline phase and then every two weeks within the intervention phase. The study schedule outlines the completion dates for each outcome measure collected (Table 2).

**Table 2:** Study schedule outlining time periods for completion of outcome measures.

| Protocol Activity | Baseline Phase | During Intervention Phase | Completion of Intervention Phase | End of follow-up (F/U) |
| --- | --- | --- | --- | --- |
|  | Days 1-5<br>(or 8 or 11) | Day 6 - 48<br>(or 9 – 51 or 12 – 54) | Days 20, 34 & 48<br>or 23, 37 & 51<br>or 26, 40 & 54 | End of week 12<br>post-intervention<br>phase |
| **Primary outcomes / Target behaviours** |  |  |  |  |
| NPRS | X (daily) | X (daily) | X | X |
| Confidence | X (daily) | X (daily) | X | X |
| Adverse events |  | X (end of week 1) | X | X |
| **Secondary outcomes / Generalization measures** |  |  |  |  |
| NDI | X |  | X | X |
| DASS-21 | X |  | X | X |
| SF12 | X |  | X | X |
| PCS | X |  | X | X |
| Global Rating of Change |  |  | X | X |

**Legend:** NPRS = numerical pain rating scale; NDI = neck disability index; DASS-21 = The depression & anxiety, stress scale - 21 items; SF12 = 12-item short form survey; PCS = pain catastrophizing scale

### Primary Outcomes – Target Behaviours

1. Individual pain experiences were measured using the PEG scale, which measures average neck pain intensity (P), how much pain interferes with enjoyment of life (E), and how much pain interferes with general activity (G), all during the last 24 hours, each measured on a 0 to 10 numeric rating scale (NRS) (Krebs et al., 2009). The standard of error is 1.8 (Krebs et al., 2010).
2. Self-efficacy refers to an individual’s belief in their capacity to execute behaviours necessary to produce specific performance attainments (Bandura et al., 1977). Confidence in performing daily activities was evaluated using the question “How confident are you in your ability to perform your daily tasks in the presence of your neck pain or disability?” with 1 indicating not at all confident; 2 indicating a little confident; 3 indicating moderately confident; 4 indicating very confident and 5 indicating extremely confident. This question has previously been used to evaluate self-efficacy in chronic WAD (Ritchie et al., 2021).

### Secondary Outcomes – Generalization Measures

Generalization measures were collected to determine whether intervention effects generalized to other behaviours and functional outcomes associated with the heterogeneity of chronic WAD, and if the effects of the intervention were sustainable. They were collected on admission to the study, and thus prior to commencing PRP; at the end of the intervention phase and then three months following interventions (Table 2). Measures were not collected after the basement phase, as it was not anticipated that changes would occur within 11-days of receiving PRP. No scheduled treatment was provided following trial completion, although participants were free to pursue treatment as they desired, which they tracked in their diaries. Measures evaluated the following:

1. Global rating of change (-5 (vastly worse) to +5 (completely recovered) 0 = unchanged) numerical rating scale) (Hurst & Bolton, 2004) with a change of ≥ 2 indicating a clinically meaningful change (Kamper, 2009). *Pain and Disability -*
2. Numeric Pain Rating Scale (NPRS) with scores ≥ 3 indicating moderate to severe neck pain and a change ≥ 2 representing a minimum clinically important difference (MCID) (Young et al., 2019).
3. Neck Disability Index (NDI; percentage score calculated) measured pain-related disability with scores ≥ 30% indicative of moderate -to-severe disability status and a change ≥ 10% representing MCID (Vernon & Mior, 1991). *Psychological Health -*
4. The Depression & Anxiety Stress Scale - 21 items (DASS-21) is a set of three self-report scales designed to measure the emotional states of depression, anxiety and stress (Lovibond & Lovibond, 1995). Normal scores for depression are < 10; anxiety < 8 and stress < 14 respectively;
5. The Pain Catastrophizing Scale (PCS) measures catastrophic thoughts related to pain with scores > 23 indicative of elevated pain catastrophizing and MCID > 6 (Sullivan et al., 1995);
6. Generic measures of health status (SF-12) (Ware et al., 1996) were collected to measure quality of both physical and mental health. Norm-based scoring applies with scores above and below 50 above and below the average of the general U.S. population. As the standard deviation is 10 for both scales, scores below 40 are indicative of less than normal quality of life. MCID in low back pain > 3 has been reported (Diaz-Arribas et al., 2017).

### Data Analysis

Each participant’s data for both target behaviours (PEG and Confidence) were graphed and analyzed at the individual level using recommended structured visual analysis (Lane & Gast, 2014) in conjunction with statistical analysis. Evaluation of the visual analyses across participants helps determine if changes measured are reliably and consistently replicated. Within-phase data patterns were evaluated individually using, (1) level, the relative change in data (between the median of the first half of the observations and the median of the last half of the observations) within each of the baseline and intervention phases; (2) variability, the range or variance of the data points around the best-fitting line with stability indicated if 80% of the data points were within 25% of the median value within each phase (Kratochwill et al., 2013; Lane & Gast, 2014). Consistency of similar phases (e.g. baseline and intervention phases) across participants was measured using the CONsistency of DAta Patterns (CONDAP) approach with the interpretation of consistency indicated as very high (0-0.5), high (0.6-1), medium (1.1-1.5), low (1.6-2) or very low (> 2.1).

Evaluation of between-phase patterns included trend – the systematic increase or decrease of data over time and assessed using a split-middle approach (Kazdin, 2011) with a positive trend indicating if the change in levels was greater or equal to MCID. The immediacy of effect was not evaluated, as the biological therapy provided (PRP) involves possible latency of change which may or may not have interacted with intervention as time progressed. Tau-U was used to determine the overlap of data points between phases. Tau-U provides an intervention effect size (ES) and a weighted average across tiers (Heyvaert et al., 2015; Parker et al., 2011). Tau-U analyses were performed using the website: http://singlecaseresearch.org. Effect size results were based on recommendations from Vannest and Ninci (Vannest & Ninci, 2015): small effect < .02, moderate effect (0.2 to 0.6), large effect (0.6 to 0.8) and large to very large effects (> 0.8).

## Results

### Participants

The six participants (mean (SD): 43.1 (9.3); 50% female) reported the presence of WAD for a median of 23 months (range: 14-68 months) at entry into the study (Table 3). Both treatment allocations were evenly matched for mean age (UC: 43 years; PT: 43.3 years) and duration of symptoms (UC: 32.3 months; PT: 31.3 months). Three participants had PRP at C2/3; two at C3/4, two at C5/6 and two at C6/7. Five participants had multiple levels injected, whilst one participant had bilateral injections. The only deviation from protocol involved admission of a participant > 55 years old. This resulted from lagging recruitment rates and to balance the age and duration of symptoms for each intervention. One participant in UC was lost to follow-up and did not complete follow-up measures 3-months following the intervention phase.

**Table 3:** Demographic data, pre-and post-generalization measures. Time: 1 = prior to PRP; 2 = post-PRP treatment (end of phase B post-intervention); 3 = 3-month follow-up.

|  | Group 1 (Treat as Usual: UC) |  |  |  |  |  |  |  |  | Group 2 (Active Intervention: PT) |  |  |  |  |  |  |  |  |
| --- | --- | --- | --- | --- | --- | --- | --- | --- | --- | --- | --- | --- | --- | --- | --- | --- | --- | --- |
|  | P1 |  |  | P2 |  |  | P3 |  |  | P4 |  |  | P5 |  |  | P6 |  |  |
| Sex | M |  |  | M |  |  | F |  |  | F |  |  | F |  |  | M |  |  |
| Age (5 years range) | 46-50 |  |  | 46-50 |  |  | 31-35 |  |  | 31-35 |  |  | 31-35 |  |  | 55-60 |  |  |
| Duration (months) | 15 |  |  | 14 |  |  | 68 |  |  | 48 |  |  | 21 |  |  | 25 |  |  |
| Baseline period (days) | 5 |  |  | 8 |  |  | 11 |  |  | 5 |  |  | 8 |  |  | 11 |  |  |
| Time | T1 | T2 | T3 | T1 | T2 | T3 | T1 | T2 | T3 | T1 | T2 | T3 | T1 | T2 | T3 | T1 | T2 | T3 |
| PEG (/30) | 14 | 15 | 1 | 23 | 13 | 9 | 19 | 15 | - | 18 | 6 | 3 | 25 | 3 | 5 | 18 | 21 | 20 |
| Confidence (1 to 5) | 3 | 2 | 4 | 2 | 2 | 3 | 3 | 3 | - | 3 | 4 | 4 | 2 | 4 | 3 | 3 | 3 | 1 |
| NRS (/10) | 4 | 5 | 0 | 7 | 3 | 2 | 6 | 7 | - | 6 | 2 | 1 | 7 | 1 | 2 | 6 | 5 | 6 |
| NDI (%) | 38 | 36 | 8 | 62 | 50 | 44 | 50 | 50 | - | 34 | 29 | 18 | 44 | 9 | 22 | 43 | 42 | 48 |
| DASS-D (/42) | 6 | 2 | 0 | 22 | 22 | 14 | 10 | 6 | - | 6 | 2 | 0 | 2 | 0 | 4 | 18 | 8 | 10 |
| DASS-A (/42) | 0 | 0 | 0 | 4 | 2 | 0 | 0 | 0 | - | 14 | 6 | 8 | 2 | 0 | 0 | 4 | 0 | 0 |
| DASS-S (/42) | 4 | 4 | 0 | 30 | 22 | 18 | 2 | 0 | - | 14 | 10 | 14 | 6 | 2 | 10 | 12 | 8 | 12 |
| PCS (/52) | 11 | 10 | 2 | 19 | 23 | 20 | 9 | 7 | - | 10 | 8 | 9 | 10 | 3 | 0 | 26 | 17 | 18 |
| SF12 physical | 38 | 35 | 40 | 35 | 30 | 30 | 41 | 32 | - | 33 | 48 | 40 | 21 | 43 | 50 | 36 | 33 | 34 |
| SF12 mental | 44 | 56 | 64 | 29 | 43 | 50 | 40 | 58 | - | 54 | 50 | 58 | 62 | 60 | 48 | 31 | 39 | 37 |
| GROC (-5 to 5) | - | 0 | 4 | - | 2 | 2 | - | 1 | - | - | 2 | 4 | - | 4 | 3 | - | 2 | 0 |

**Legend:** PEG: Pain Intensity (Numeric Rating Scale/10); Pain Interference with Enjoyment of Life (Numeric Rating Scale/10) and Pain Interference with General Activity (Numeric Rating Scale/10); NRS: Numerical Rating Scale; GROC: Global Rating of Change; NDI: Neck Disability Index; DASS: Depression, Anxiety and Stress scale; PCS: Pain Catastrophization Scale; SF12: Short Form12.

### Treatment Fidelity/Adherence

For PT, one participant (P4) only attended two of the scheduled six sessions; one participant (P5) attended six sessions and one participant attended seven sessions. None of the participants recorded the frequency of home exercise completion in their daily diaries. Following the intervention phase, one participant (P4) attended three further physiotherapy sessions in the ensuing 3-month follow-up phase; one (P5) attended four sessions, and one (P6) attended seven chiropractic sessions.

In UC, two participants (P1 and P2) accessed the study physician for advice on reducing pain. One participant contacted the physician twice and one participant on a solitary occasion. One participant (P2) completed 31 days of exercises contained within the take home booklet. The participant also attended one massage therapy appointment. The other two participants did not record their exercise frequency. During the ensuing 3-month follow-up period, one participant (P1) attended eight sessions of physiotherapy and two sessions of massage therapy. One participant (P2) attended 13 sessions of physiotherapy, two sessions of massage therapy and received greater occipital nerve blocks for headache management (which did not result in any change in symptoms). The other participant was lost to follow-up.

### Primary Outcome Measures/Target Behaviours

#### PEG

For UC, visual analysis of daily PEG showed a stable baseline for P1 and P2, with a reduction in PEG for P3 (Figure 1). P2 and P3 demonstrated an improvement in PEG from the baseline to intervention phase (i.e. trend) with a low level of consistency (i.e. CONDAP). Statistically, the three participants demonstrated a significant reduction in PEG scores with large (P3) to very large (P1 and P2) effect sizes demonstrated. There was a significant weighted Tau-U across tiers (Z = -3.30; p < .001; ES = -.75 (90%CI: -.53 to -.98) (Table 3).

For PT, two participants demonstrated improvements in the baseline phase (P4 and P5), with concurrent increased variability in their PEG scores. Visual analysis only showed a trend for improvement from the baseline to intervention phase for P5, although there was very low consistency demonstrated (i.e. CONDAP) (Figure 1). However, statistical analysis demonstrated a significant reduction in PEG scores for all three participants with very large effect sizes demonstrated. As per UC, there was a significant weighted Tau-U across tiers (Z = -3.57; p < .001; ES = -.82 (90%CI: -.59 to -1.0) (Table 3). The overlapping confidence intervals demonstrated that there was no significant difference between treatment allocations in PEG scores.

### Confidence in Daily Tasks

Two of the participants in each treatment allocation improved during the baseline phase for confidence in performing their daily tasks in the presence of their neck pain (Appendix Fig. 1). However, there was significant variability in their observations during this phase. Only P2 in UC, and P5 and P6 in PT demonstrated statistically significant improvement. This was likely due to significant variability in observations during the intervention phases with only P3 demonstrating stability of measures during this phase. There was also medium to low consistency across tiers during the intervention phase. However, weighted TAU-U was significant for both treatment allocations (UC: Z = 2.18; p < .001; ES = .50 (90%CI: .28-.72); PT: Z = 1.93; p = .0012; ES = .46 (90%CI: .22-.69). Again, overlapping confidence intervals showed that there were no significant differences in the change of confidence between phases for either treatment allocation.

### Secondary Outcome Measures/Generalization Measures

#### Global Rating of Change

One person in UC (P2) reported improvement exceeding MCID (≥ 2) following the intervention phase. All three people in PT exceeded MCID following the intervention phase. After the subsequent 3-month follow-up phase, two persons in UC (1 missing data) and two of the three participants in PT reported improvement exceeding MCID. No persons reported a worsening of condition at any stage of follow-up.

### Pain and Disability

At baseline, all six participants reported moderate-to-severe levels of disability (NDI ≥ 30%) and moderate to high levels of pain (NRS ≥ 3). Following the intervention phase, one person in UC (P2) and one person in PT (P5) demonstrated improvement exceeding MCID (≥ 10%). Three months following PRP, two persons in each treatment allocation exceeded MCID (≥ 10%), with one person in UC and two persons in PT improving to a mild disability level (NDI < 30%). For pain, one person in UC (P2) and two persons in PT (P4 and P5) reported significant improvements in pain (> 2) following the intervention phase. Two persons in each treatment allocation exceeded MCID for pain after a further 3-months follow-up phase.

### Psychological Health

At baseline, one participant (P2) reported high depressive and stress symptoms; one participant (P4) high anxiety symptoms and one participant (P6) high depressive symptoms and high levels of pain catastrophization. Both P4 and P6 reported normal levels of symptoms following the intervention phase, although after the follow-up phase, P4 reported recurrence of anxiety symptoms and P6 reported depressive symptoms (with ongoing improvement in pain catastrophization). P2 continued to report high levels of depressive and stress symptoms.

Five participants (excepting P3) reported significantly lower levels of physical health (SF12 physical < 40) at baseline when compared to the normal population. Two participants (P2 and P6) reported lower levels of mental health (SF12 mental < 40). Two of the participants (P4 and P5) in PT improved physical health within population norms following the intervention phase, which were maintained during the follow-up phase. All three participants in UC reported ongoing reduced levels of physical health (i.e. P3 worsened) following the intervention and follow-up phases. Improvements in mental health were reported for P2 following UC, which was maintained during the follow-up phase. P6 reported improved mental health scores (> 3) following the intervention phase, however it was still not within normal population limits.

## Discussion

This is the first trial to evaluate the effects of rehabilitation in people with chronic WAD receiving cervical facet joint PRP injections. All six participants demonstrated a statistically (Tau-U) significant reduction in PEG scores following PRP, irrespective of intervention received. In addition, three of the six participants demonstrated a significant improvement in confidence to complete daily tasks in the presence of neck pain. In concordance with the primary outcomes, generalization measures demonstrated that four of the six participants reported a significant improvement in their recovery from whiplash injuries following the intervention phase, with four participants also reporting a significant improvement in their recovery 3-months later (GROC). Three participants significantly improved pain levels following the intervention phase, with only one participant reporting moderate-to-severe pain after the ensuing follow-up period. Similarly, significant improvements were reported for disability. Two participants reported improvement following the intervention phase, whilst three of the five participants reported no or mild disability 3-months later. For the three participants with initial depressive symptomatology, two reported improvement following the intervention phase, whilst the one person with anxiety symptoms also reported improvement following this phase. The participant with elevated stress symptoms was unchanged after the intervention phase. Physical health quality of life (SF12 physical) improved to normal population limits in two of the six participants following the intervention phase and in three of the five participants after the follow-up period. Mental health quality of life (SF12 mental) significantly improved for both participants with initial scores below population norms. Overall, five of the six participants reported mental health scores within population norms after the intervention phase. Thus, these individual improvements showed the utility of SCED methodology, which would not have been possible with study designs focused on group-level evaluation.

Our study, utilizing a SCED demonstrated the possible utility of physiotherapy in improving outcomes at the individual level, which contrasts from group-level analyses generally reported in the literature, whereby it has been shown that physiotherapy interventions provide modest benefits when compared to control interventions (Jull et al., 2007; Ludvigsson et al., 2015; Michaleff et al., 2014; Ris et al., 2016; Stewart et al., 2007; Sutton et al., 2014). Individually, two of the three participants receiving PT reported a significant reduction in PEG, improved confidence in performing activities in pain, reduced neck pain intensity and one of these participants reported a significant reduction in disability, whilst the other participant attended further physiotherapy following the intervention phase and reported a significant improvement in disability 3-months later. Two of the three PT participants also improved their physical health quality of life by the completion of the intervention phase. None in UC improved their physical health quality of life by this stage. The one participant in PT with reduced mental health quality of life also significantly improved by this stage, albeit not quite reaching normative population values. Only one in UC reported a reduction of neck pain at the completion of the intervention phase. Both participants in UC (who completed follow-up questionnaires 3-months later) attended subsequent physiotherapy (eight or 13 sessions respectively) during the 3-month follow-up phase (following UC) and reported a significant improvement in pain, disability and mental health quality of life, whilst one also reported that their physical health quality of life was within population norms, indicating the possible utility of physiotherapy when evaluated at the individual level.

Although this study was not designed to investigate group differences, overlapping confidence intervals from aggregated individual responses suggested that there were no differences in health outcomes between PT and UC. There are several possible reasons. Previous studies have enrolled participants for physiotherapy programs anywhere from 6-16 weeks (Jull et al., 2007; Ludvigsson et al., 2015; Michaleff et al., 2014; Ris et al., 2016; Stewart et al., 2007). Thus, an appropriate dose response to physiotherapy may not have been fully observed in the six-weeks we chose to pursue in this study. Our data suggests that individuals continued to improve over the following 3-months – for pain and disability, physical and mental health. Thus, future studies may wish to increase the duration of physiotherapy to further improve outcomes. The type of physiotherapy provided in this study was primarily focused on previously published treatment directed towards individual physical impairments via progressive low load, motor control, strengthening and endurance exercises (Jull et al., 2018; Ludvigsson et al., 2015). Physical activity guidelines were also provided, although a recent study demonstrated that there was no improvement in health outcomes for people with chronic WAD who were prescribed physical activity (Ludvigsson et al., 2015). Although general advice and reassurance was provided, no specific psychologically informed PT was provided that may have addressed personal psychological factors associated with an individual’s pain experience (Campbell et al., 2018; Carroll et al., 2006; Sterling et al., 2019). However, it is unclear if this would have significantly improved the primary outcomes – particularly PEG scores, as the addition of a behavioural approach has not resulted in reductions in pain intensity for people with chronic WAD (Ludvigsson et al., 2015; Soderlund & Lindberg, 2001; Wicksell et al., 2008), although psychological outcomes have been shown to improve with interdisciplinary therapy (Soderlund & Lindberg, 2001; Wicksell et al., 2008). Interestingly, of those with above-threshold psychological symptom profiles at baseline in our study, only one participant (P6) demonstrated persistent moderate to severe pain and disability levels at study completion. A recent study demonstrated that trauma-focused cognitive behavioural therapy did not improve health outcomes more than supportive therapy (psycho-education, problem solving and diary completion of mood states) in chronic WAD for people with post-traumatic stress disorder (Andersen et al., 2021). It may be that the group receiving UC, which involved provision of a booklet with evidence informed psycho-education, together with the ability to speak to study personnel over a 6-week period may have resulted in a form of ‘supportive therapy’ that assisted with improvements in health outcomes. Thus, further research is warranted to determine what optimal treatment is for chronic WAD.

Lack of improvement in the PT group may also be due to treatment adherence. Participants in PT did not attend all their pre-scheduled appointments. It is also unclear whether they were diligent in adherence to the home exercises provided. Although participants were provided with individualized exercise and physical activity guidelines, none receiving PT reported the frequency of home exercise participation in their daily diaries. Given that one of the UC participants reported in their diary that they exercised on 31 of the 42 days, it may be that the home exercise component was crucial to improvement, irrespective of manual therapy and dry needling which have not previously demonstrated improvements in outcomes in chronic WAD (Gross et al., 2007; Gross et al., 2002; Sterling et al., 2015). Finally, although sample size recommendations suggest that a minimum of five observations are required for each person to demonstrate significant differences at an individual level (Romeiser Logan et al., 2008), this proof-of-concept study was underpowered to demonstrate group differences when results were aggregated. This is especially pertinent for interventions with lack of immediacy of effect (such as rehabilitation) when considerable variability in daily target behaviours (e.g. pain) occur as time progresses. This was evidenced by the very low levels of confidence demonstrated in both interventions across tiers, both in the baseline and intervention phases. This study provides direction moving forwards, with precision estimates demonstrating an effect size favouring PT of .07.

This is the first trial to demonstrate the effect of PT or UC following PRP in chronic WAD and cervical facet joint mediated pain. This trial demonstrated that all participants, irrespective of intervention allocation, showed large to very large effect sizes for both primary outcomes – reduction in PEG and confidence in performing daily activities with neck pain. UC or PT following PRP also resulted in improvements in physical and mental health. Although there wasn’t a high psychological symptom profile burden in participants, it is also noteworthy that individuals improved both physical and psychological health outcomes. The model of facet joint pain in WAD is well established (Curatolo et al., 2011). In vivo models of painful cervical facet joint injury have demonstrated that a complex interaction of electrophysiological, inflammatory and nociceptive signaling cascades are associated with whiplash facet joint pain (Dong et al., 2008; Dong et al., 2012; Kras et al., 2014; Quinn et al., 2010; Quinn & Winkelstein, 2011). Thus, the improvements demonstrated by PRP may have been due to modulation of any or all of these underlying processes. The exact mechanism of action for PRP is unknown, but likely augments multiple aspects of tissue repair. PRP contains supraphysiologic levels of growth factors and cytokines (Amable et al., 2013). The alpha granules in platelets also secrete growth factors that are essential for tissue repair (Sampson et al., 2008). The growth factors also increase collagen content, promote endothelial regeneration, and stimulate angiogenesis (Podd, 2012; Sampson et al., 2008). In this trial, PRP demonstrated significant improvement in PEG scores in the basement phase in 50% of participants. Given that significant immediate reductions in pain occurred, it would suggest that PRP demonstrated an anti-nociceptive effect, as distinct from a tissue repair effect which would have required greater than the initial 5-11 days basement phase for improvements in outcomes to be demonstrated. A recent systematic review found that raised blood inflammatory markers (interleukin 1β and tumour necrosis factor α) were present in chronic neck pain and that these were associated with pain and disability measures (Farrell et al., 2020). Thus, it is plausible that those demonstrating immediate improvements in this trial presented with underlying inflammatory mechanisms which were associated with their clinical symptoms. However, the maintained benefits evident 4-5 months post-PRP may also be indicative of improvements in collagen composition (Podd, 2012). It is possible that physiotherapy may assist with this aspect of rehabilitation by gradually increasing load on the capsular tissue through appropriate goal-directed exercises. It must be noted that PRP delivered to lumbar spine facet joints demonstrated improvements in clinical outcomes over time and possibly without rehabilitation (Wu et al., 2017), suggesting that the effect of rehabilitation cannot yet be fully determined.

There are several limitations with this trial that require attention. In particular, there was a lack of adherence to anticipated and prescribed treatment for PT. However, all participants in PT demonstrated significant improvements in PEG and two of the three demonstrated improvements in confidence to perform activities when in pain, irrespective of their attendance or diligence to exercise prescription. We were also unable to blind the participant and practitioner to the phase of the study. Assessors were also not blind to all phases of the study, although they were independent. This study was also underpowered for group comparisons, although as a proof-of-concept study, we were able to establish the effect size of physiotherapy for future trials aggregating results across tiers, especially if repeated observations have higher variability within phases or there is low consistency across tiers as was apparent in this trial. Although there was no cost for physiotherapy in this trial, participants paid for PRP. It is well recognized that there is a positive association between wealth and better health (Braveman et al., 2010). Thus, the improved treatment outcomes may reflect other factors that may not generalize to the overall population.

Our study also provides preliminary data on the effect of PRP in chronic WAD in a highly-selected sample of people with an underlying homogeneous site of anatomical nociception. This study followed participants for between four to five months. Longer term effects of PRP need to be reported to determine dose responsiveness. Cost effectiveness also needs to be determined over longer time periods, so an accurate comparison against other therapeutic modalities are possible - e.g. neck specific exercises (Ludvigsson et al., 2015) or 12-month RFC outcomes (Lord et al., 1996).

## Conclusion

Physiotherapy or usual care following PRP resulted in significant improvements in pain intensity, pain interference with enjoyment of life and general activity and improvements in confidence to perform activities of daily living with neck pain. Other health outcomes reinforced these findings, supporting the premise that these results could be generalized.

## Data Availability

All data produced in the present study are available upon reasonable request to the authors

## Acknowledgements

The authors would like to acknowledge EFW Radiology and Evidence Sport & Spinal Therapy for assisting with participant recruitment and providing material support (PRP kits). The authors would like to thank Dr. Chirag Patel and Dr. Rob Burnham for their contributions to the provision of PRP in this study.

