## Supplemental Table 1 for "The Efficacy of Multimodal Physiotherapy versus Usual Care in Chronic Whiplash-Associated Disorders With Facet-Mediated Pain Undergoing Platelet Rich Plasma treatment: A Series of Single Case Experimental Designs"

**Supplementary Table 1:** Interventions delivered to study participants

*Platelet-Rich Plasma (PRP)*

PRP was prepared by a process known as differential centrifugation. This was completed using the Harvest SmartPRep 2 APC+, which is a standardized commercially available kit. PRP was obtained from a sample of participants’ blood drawn at the time of treatment. A 30cc venous blood draw yields 3-5cc of PRP. The blood draw occurs with the addition of an anticoagulant, acid citrate dextrose to prevent platelet activation prior to its use. A two-spin centrifugation is completed. The initial centrifugation is to separate red blood cells (RBC), followed by a second centrifugation to concentrate platelets, which are suspended in the smallest final plasma volume. The preparation was injected using a sterile technique with fluoroscopic imaged guidance.

Interventional Procedure

The patient was appropriately consented for the procedures. The patient was placed prone on the procedure table. The skin was then prepared and sterilized. There was no local anesthetic injected to prevent premature platelet damage. The needle was positioned into the intra-articular facet joint. Omnipaque 300 was used to confirm position and a facet arthrogram was obtained. Once satisfactory, 1.25 cc of the PRP was injected into each joint.

*Educational Pamphlet*

All participants were provided with an educational pamphlet on PRP, providing an outline of the treatment received, together with possible risks and precautions. This pamphlet also provided advice on appropriate self-care following PRP, including use of heat and possible medications that could be used for symptomatic relief of residual post-treatment soreness.

Group 1: *Usual Care/Informational Booklet*

Individuals randomized to usual care (UC) were provided with an advice booklet *Whiplash Injury Recovery: A Self Help Guide (3^rd^ edition)*^29^. It provides information about whiplash; assurance about prognosis; advice to stay active and resume working as well as information on correct posture; pictorial descriptions of specific exercises for the neck and upper limbs and information on resuming functional daily activities. The booklet is based on the recommendations of the current Australian Guidelines for Whiplash Management^30^. To assist with balancing time requirements between treatment allocations, UC included the option to attend weekly review appointments (which were completed on the telephone if desired) with a medical doctor; primarily to review the information in the booklet and progress the ‘general’ exercises and activity recommendations within the booklet. No in-person physiotherapy was provided. Daily primary outcome measures were collected during the six-week period. Participants were routinely followed up at the end of the six-week period with a medical doctor.

Group 2: *Multimodal Physiotherapy*

Participants were offered a maximum of two sessions in weeks 1-4 and one session in weeks 5 and 6. Participants were responsible for booking or cancelling the pre-scheduled appointments, depending on their individual needs. The exercise program comprised specific tailored exercises for each participant to improve the movement and control of the neck and shoulder girdle, which have previously been demonstrated to be altered in chronic WAD. The program started with a clinical examination of the cervical muscles and the axio-scapular-girdle muscles and included tests that assess ability to recruit the muscles in a coordinated manner, and tests of muscle endurance at low levels of maximum voluntary contraction. The specific impairments that were identified were then addressed with an exercise program that was supervised and progressed by the physiotherapist. This specific treatment program has been described in detail^22, 34^ and focuses on activating and improving the coordination and endurance capacity of the neck flexor, extensor and scapular muscles in specific exercises and functional tasks. The exercise program follows the current Australian guidelines for the management of chronic whiplash^4^. The exercises were of a low load nature and designed to be pain free. At the same time, the physiotherapist guided the participant’s return to normal activities. Participants were also instructed to perform the exercises at home, once per day. Written and illustrated exercise instructions were provided. An electronic spreadsheet was provided to participants to record compliance with the exercises. Although participants were encouraged to use this spreadsheet and track their goals, this feature was not formally checked unless requested by the participant.

*Co-interventions:*

On the day of PRP, the interventional physician recommended use of over-the-counter medication for procedural related pain. NSAID medications were prohibited in the 14 days post-PRP due to their possible inhibitory effect on PRP. The criteria for recommending medication was based on worsening pain that was debilitating in nature (in the short term) or continuing high levels of pain that had not improved after the first week of treatment. Participants who experienced high levels of continuing or worsening pain were able to contact research staff to access the study doctor for advice over the phone and possible return for an earlier review with the physiotherapist. No participant contacted the doctor or attended the physiotherapist over the study period for procedural pain.
