## Supplemental Figure 1 for "The Efficacy of Multimodal Physiotherapy versus Usual Care in Chronic Whiplash-Associated Disorders With Facet-Mediated Pain Undergoing Platelet Rich Plasma treatment: A Series of Single Case Experimental Designs"

**Appendix:**

**Supplementary Figure 1:** A two section AB graph plotting confidence scores of six people undergoing PRP.

**Legend: Confidence** in performing activities of daily living in the presence of neck pain for phases A (baseline = left of vertical line) and B (intervention) and shown for Group 1 = Usual Care (P1, P2, P3) and Group 2 = Physiotherapy (P4, P5, P6) participants.

|  | P1 |
| --- | --- |
| 1 | 2 |
| 2 | 2 |
| 3 | 2 |
| 4 | 3 |
| 5 | 3 |
| 6 | 3 |
| 7 | 3 |
| 8 | 3 |
| 9 | 3 |
| 10 | 3 |
| 11 | 3 |
| 12 | 2 |
| 13 | 3 |
| 14 | 2 |
| 15 | 2 |
| 16 | 2 |
| 17 | 3 |
| 18 | 3 |
| 19 | 3 |
| 20 | 3 |
| 21 | 3 |
| 22 | 3 |
| 23 | 3 |
| 24 | 3 |
| 25 | 3 |
| 26 | 3 |
| 27 | 4 |
| 28 | 4 |
| 29 | 3 |
| 30 | 3 |
| 31 | 3 |
| 32 | 2 |
| 33 | 3 |
| 34 | 3 |
| 35 | 3 |
| 36 | 3 |
| 37 | 2 |
| 38 | 2 |
| 39 | 2 |
| 40 | 2 |
| 41 | 3 |
| 42 | 2 |
| 43 | 2 |
| 44 | 2 |
| 45 | 2 |
| 46 | 3 |
| 47 | 2 |

P2

|  | P2 |
| --- | --- |
| 1 | 1 |
| 2 | 1 |
| 3 | 1 |
| 4 | 1 |
| 5 | 1 |
| 6 | 1 |
| 7 | 1 |
| 8 | 1 |
| 9 | 2 |
| 10 | 2 |
| 11 | 2 |
| 12 | 2 |
| 13 | 2 |
| 14 | 2 |
| 15 | 2 |
| 16 | 2 |
| 17 | 2 |
| 18 | 2 |
| 19 | 2 |
| 20 | 2 |
| 21 | 2 |
| 22 | 2 |
| 23 | 2 |
| 24 | 2 |
| 25 | 3 |
| 26 | 3 |
| 27 | 3 |
| 28 | 3 |
| 29 | 3 |
| 30 | 3 |
| 31 | 2 |
| 32 | 2 |
| 33 | 3 |
| 34 | 3 |
| 35 | 3 |
| 36 | 3 |
| 37 | 3 |
| 38 | 1 |
| 39 | 2 |
| 40 | 2 |
| 41 | 2 |
| 42 | 1 |
| 45 | 3 |
| 46 | 3 |
| 47 | 3 |
| 48 | 3 |
| 49 | 3 |
| 50 | 3 |

P3

|  | P3 |
| --- | --- |
| 1 | 4 |
| 2 | 2 |
| 3 | 2 |
| 4 | 2 |
| 5 | 2 |
| 6 | 3 |
| 7 | 3 |
| 8 | 4 |
| 9 | 4 |
| 10 | 4 |
| 11 | 4 |
| 12 | 4 |
| 13 | 5 |
| 14 | 5 |
| 15 | 5 |
| 16 | 5 |
| 17 | 3 |
| 18 | 4 |
| 19 | 3 |
| 20 | 3 |
| 21 | 3 |
| 22 | 3 |
| 23 | 2 |
| 24 | 3 |
| 25 | 4 |
| 26 | 4 |
| 27 | 4 |
| 30 | 4 |
| 31 | 4 |
| 32 | 2 |
| 33 | 2 |
| 34 | 4 |
| 35 | 2 |
| 36 | 3 |
| 37 | 4 |
| 38 | 4 |
| 39 | 4 |
| 40 | 4 |
| 41 | 3 |
| 42 | 4 |
| 43 | 4 |
| 44 | 4 |
| 45 | 4 |
| 46 | 4 |
| 47 | 3 |
| 48 | 3 |
| 49 | 4 |
| 50 | 4 |
| 51 | 4 |
| 52 | 4 |
| 53 | 4 |

|  | P4 |
| --- | --- |
| 1 | 3 |
| 2 | 3 |
| 3 | 3 |
| 4 | 4 |
| 5 | 4 |
| 6 | 3 |
| 7 | 3 |
| 8 | 3 |
| 9 | 3 |
| 10 | 3 |
| 11 | 3 |
| 12 | 3 |
| 13 | 2 |
| 14 | 2 |
| 15 | 3 |
| 17 | 2 |
| 18 | 2 |
| 19 | 2 |
| 20 | 3 |
| 21 | 4 |
| 22 | 3 |
| 23 | 3 |
| 24 | 3 |
| 25 | 4 |
| 34 | 4 |
| 35 | 4 |
| 36 | 4 |
| 37 | 4 |
| 38 | 4 |
| 39 | 4 |
| 40 | 4 |
| 42 | 4 |
| 43 | 4 |
| 44 | 3 |
| 45 | 3 |

|  | P5 |
| --- | --- |
| 1 | 2 |
| 2 | 2 |
| 3 | 2 |
| 4 | 2 |
| 5 | 2 |
| 6 | 3 |
| 7 | 3 |
| 8 | 2 |
| 9 | 3 |
| 10 | 3 |
| 11 | 3 |
| 12 | 3 |
| 13 | 3 |
| 18 | 3 |
| 20 | 3 |
| 21 | 3 |
| 22 | 3 |
| 23 | 3 |
| 24 | 3 |
| 25 | 3 |
| 26 | 3 |
| 27 | 3 |
| 28 | 3 |
| 29 | 3 |
| 30 | 3 |
| 31 | 3 |
| 32 | 3 |
| 33 | 4 |
| 34 | 4 |
| 35 | 4 |
| 36 | 4 |
| 38 | 4 |
| 39 | 4 |
| 40 | 3 |
| 41 | 4 |
| 42 | 4 |
| 45 | 4 |
| 46 | 4 |
| 47 | 4 |
| 48 | 5 |
| 49 | 5 |
| 50 | 5 |

|  | P6 |
| --- | --- |
| 1 | 2 |
| 2 | 2 |
| 3 | 2 |
| 4 | 2 |
| 5 | 2 |
| 6 | 2 |
| 7 | 2 |
| 8 | 2 |
| 9 | 2 |
| 10 | 2 |
| 11 | 2 |
| 12 | 2 |
| 13 | 2 |
| 14 | 2 |
| 15 | 2 |
| 16 | 2 |
| 17 | 2 |
| 18 | 2 |
| 19 | 2 |
| 20 | 2 |
| 21 | 2 |
| 22 | 2 |
| 23 | 2 |
| 24 | 2 |
| 25 | 2 |
| 26 | 3 |
| 27 | 2 |
| 28 | 2 |
| 29 | 2 |
| 30 | 3 |
| 31 | 3 |
| 32 | 3 |
| 33 | 3 |
| 34 | 3 |
| 35 | 2 |
| 36 | 3 |
| 37 | 3 |
| 38 | 3 |
| 39 | 3 |
| 40 | 3 |
| 41 | 3 |
| 42 | 3 |
| 43 | 3 |
| 44 | 3 |
| 45 | 3 |
| 47 | 3 |
| 48 | 3 |
| 49 | 3 |
| 50 | 3 |
| 51 | 3 |
| 52 | 3 |
| 53 | 3 |
